# Conjunctivitis outbreak caused by Enterovirus Type C in Luzira prisons, Uganda, February–April 2024

**DOI:** 10.1101/2025.04.04.25325059

**Authors:** Hannington Katumba, Richard Migisha, Charity Mutesi, Emmanuel Mfitundinda, Joanita Nalwanga, Joyce Owens Kobusingye, Daniel Wenani, Loryndah Olive Namakula, Emmanuel Okello Okiror, Janet Lubega Kobusinge, Gertrude Abbo, Annet Mary Namusisi, Bridget Ainembabazi, Patrick Kwizera, Wilfred Opeli, Winnie Agwang, Esther Nabatta, Tracy Maureen Rutogire, Ritah Namusoosa, Samuel Lugwana, Lilian Bulage, Benon Kwesiga, Samuel Gidudu, Alex Riolexus Ario

## Abstract

**Background:** On March 7, 2024 the Ministry of Health (MoH) was notified of a rising number of inmates with suspected conjunctivitis in Luzira prisons in Uganda. We investigated this outbreak to determine its cause and extent, identify risk factors and recommend evidence-based control measures.

**Methods:** We investigated the outbreak in four prisons: Kampala Remand Prison (KRP), Murchison Bay Prison (MBP), Luzira Upper Prison (LUP) and Luzira Women Prison (LWP). A suspected case was onset of redness in one or both eyes with ≥1 of tearing, discharge, grainy sensation, itching, pain, or swelling, in any Luzira Prison resident from February 1 to April 3, 2024. We identified cases using medical records, and active case search among prisoners. We administered structured questionnaires and collected conjunctival swabs for PCR testing. We assessed movement of prisoners, handwashing, isolation practices, and administration of eye medications. We compared exposures of 200 randomly-selected cases 200 controls for the case control study. Using logistic regression, we conducted multivariable analysis to identify risk factors.

**Results:** We recorded 1,935 cases; overall attack rate was 23% (1,935/8,518), varying by prison: MBP (41%; 1,229/3,000), KRP (33%; 610/1,835), LUP (12%; 83/670), and LWP (0.4%; 13/3,013). With no associated deaths, most cases resolved within 4-5 days. Of the 10 samples tested, 4 (40%) were positive for Enterovirus Type C. Sharing of eye medication (aOR: 5.3, 95%CI: 2.8-9.9), and non-frequent handwashing increased odds of infection (aOR: 5.8, CI: 3.5-9.6). Prisoner mixing took place during a plea-bargain session amidst the outbreak. Symptomatic case-persons were isolated for 3 days.

**Conclusions:** The outbreak was caused by Enterovirus Type C, facilitated by prisoner mixing, sharing eye medications, short isolation periods, and inadequate hand hygiene practices. Improving infection prevention and control measures, including strict isolation, individualized eye medication, and enhanced hand hygiene practices, could prevent future outbreaks in similar settings.

## Introduction

Conjunctivitis, characterized by inflammation of the ocular mucous membranes, arises from diverse aetiologies, including viral, bacterial, allergic, parasitic, and non-specific causes (1, 2). Viruses account for approximately 80% of acute cases, with outbreaks predominantly associated with adenoviruses and enteroviruses, particularly Enterovirus type D (EV70) and Enterovirus type C (CVA24), the causative agents of acute haemorrhagic conjunctivitis (AHC) (3). The disease, has a short incubation period of 12 to 48 hours(4). AHC is highly contagious, spreading rapidly among persons through direct contact, or indirectly through sharing of beddings, clothes, eye glasses, and eye medications. Outbreaks are more common in congested settings such as schools, military barracks, prisons, and refugee camps (5, 6).

In Uganda, the first reported outbreak of conjunctivitis was in 2010 in 26 districts followed by another outbreak that occurred in Gulu District in November 2016 among inmates in a prison. Both outbreaks were caused by Coxsackievirus CVA24 (7–9). Other viruses associated with these outbreaks include Enterovirus-70, and human adenovirus species (10–13). The rapid transmission of these viruses is further facilitated by factors such as elevated temperatures, overcrowding, and high humidity (6, 14).

In February 2024, prisoners with conjunctivitis were remanded at Kampala Remand Prison in Luzira, after being transferred from a police post within Kampala City. The Kampala Capital City Authority (KCCA) notified the Ministry of Health (MoH) on March 7, 2024, about a rising number of conjunctivitis cases (n=314) within Luzira Prisons. We conducted an investigation to identify the cause and scope of the outbreak, assess the risk factors contributing to its spread, and recommend evidence-based measures to control the outbreak and prevent future similar outbreaks.

## Methods

### Outbreak setting

The outbreak occurred in Luzira Prisons located in Nakawa Division in Kampala, Uganda. It is a prison for both males and females. It is a complex of four different prisons: Luzira Women Prison, Murchison Bay Prison, Luzira Upper Prison, and Kampala Remand Prison. Initially designed to accommodate 1,700 inmates, the prison had a population of nearly 8,000 inmates at the time of the investigation. Each of the 4 prisons has a health facility at the level of health centre III.

Murchison Bay Prison is unique in that it houses the referral hospital (Murchison Bay Hospital) for all Uganda prisons, only sick inmates are admitted there. The referral hospital has a dedicated ophthalmic clinical officer, despite the ophthalmic clinic lacking basic diagnostic tools. The prison had approximately 3,000 inmates despite its intended capacity being only 600 inmates. Additionally, it had a designated isolation block, which was crowded.

KRP has a frequent prisoner traffic. Daily, about 250 inmates exit the prison to attend court sessions, some inmates are released from court, and new crime suspects are remanded. It was designed for 600 inmates; however, it was accommodating about 1,840 inmates during this outbreak. The prisoners sleep in wards, equivalent to dormitories. The prison has 12 prison wards and it houses inmates on short-term sentence, ranging from one day to 19 years.

In contrast, LUP has much less frequent in-flows of new inmates as it houses only long-term convicts serving 20 years or more. By the time of this investigation, the prison had 3,013 inmates. Luzira Women’s Prison is Uganda’s maximum-security prison for female inmates, accommodating 670 at the time of investigation.

### Case definition and case finding

We defined a suspected case as the onset of redness in one or both eyes with one or more of the following: tearing, discharge, grainy sensation, itching, pain, or swelling, in a resident at Luzira Prison from February 1, 2024, to April 3, 2024. A confirmed case was a suspected case with a positive laboratory result. We found cases systematically through review of medical records at the health facilities located within the respective prisons in Luzira. We also actively searched for cases from among inmates with the help of health workers, and the “doctor ward”, who are the equivalent of community health workers (CHWs), and generated a line list.

### Laboratory investigations

Conjunctival swabs were collected from ten suspected cases for Polymerase Chain Reaction (PCR) testing and gene sequencing at the Uganda Virus Research Institute (UVRI).

### Environmental assessment

We assessed potential factors contributing to the introduction and spread of conjunctivitis within the prisons. These included movement of prisoners, interactions between inmates from different facilities, and availability and functionality of handwashing facilities. We assessed how case-persons were managed by interviewing the case-patients, ‘doctor wards’, and health facility medical workers. We assessed for administration of eye medication and isolation of identified case-persons.

### Descriptive epidemiology

We computed attack rates (AR), stratified by prison based on the population at the time of the investigation. We described cases by clinical manifestations and likely exposures, and also constructed an epidemic curve to show the distribution of cases over time.

### Hypothesis generation

We generated hypotheses from the descriptive epidemiology and hypothesis-generating discussions with ‘doctor wards’, health facility staff, and prison officers in charge of welfare and reception, about potential risk factors for transmission of conjunctivitis. The interviews included questions on the flow of prisoners, sleeping next to someone sick, movement outside current prison, isolation of infected inmates, availability of handwashing facilities, administration of eye medication, and receiving visitors.

### Case-control study

A prison-based, case-control study was conducted. Inmates presenting with symptoms typical of conjunctivitis from Kampala Remand and Murchison Bay Prisons were enrolled in the study since these were the most affected. After administering treatment, inmates who were willing and able to provide consent were invited to participate in the interview. Signed and verbal consent were sought from the inmates after explaining the aims and procedures of the study.

Controls were selected from the same prisons as the cases. For each case, one control was selected (i.e. 1:1 ratio). The outcome of interest in our study was conjunctivitis and the primary exposure for cases and controls was contact with a case. Other exposures included; WASH habits such as washing hands with soap and water, frequency of washing hands, sharing eye ointment, and sharing personal items like blankets, sponges, and uniforms.

The sample size was estimated according to Joseph L. Fleis’s method for an unmatched case-control where from the previous study of conjunctivitis among inmates in Luzira prisons, the prevalence of exposure among controls P_0_=51% and the ratio of controls to cases considered in our study was 1:1. Total number of cases were 200 at a power of 80% and 95% confidence interval. Data were collected using interviewer-administered questionnaires that were electronically uploaded on the Kobo tool box. We identified risk factors associated with getting the infection using multivariable logistic regression.

Independent variables with a p value less than 0.05 were used to provide adjusted odds ratios with a 95% confidence interval as well as variables considered important from literature. Backward elimination was used to include variables in the final model. The most parsimonious model was selected based on the least Akaike Information Crieterion (AIC) and Bayesian Information Criterion (BIC) value.

### Ethical considerations

This study was conducted in response to a public health emergency and as such it was determined to be non-research. The MoH authorized this study and the Office of the Associate Director for Science, Center for Global Health, US Centers for Disease Control and Prevention (CDC) determined that this activity was not human subject research and with its primary intent being for public health practice or disease control. This activity was reviewed by CDC and was conducted consistent with applicable federal law and CDC policy. §§See e.g., 45 C.F.R. part 46, 21 C.F.R. part 56; 42 U.S.C. §241(d); 5 U.S.C. §552a; 44 U.S.C. §3501 et seq.

We obtained permission to investigate from the Uganda Prisons Headquarters. We obtained verbal consent from all the respondents since inmates were not allowed to hold or use pens. There were no <18year-old respondents as Luzira Prison does not detain such. ALL participants were assured that their participation was voluntary and that there would be no negative consequences for declining participation in the investigation. All cases identified during case-finding were referred to the prison health facility staff for further management. Data collected did not contain any individual personal identifiers and information was stored in password-protected computers, which were inaccessible to anyone outside the investigation team.

## Results

### Descriptive epidemiology

A total of 1,935 cases were affected by conjunctivitis in Luzira Prisons by April 2, 2024 with no deaths reported. Of these, 4 were confirmed to be caused by Enterovirus type C. The mean age of case-patients was 30 years (SD=9.4) and most of them recovered within 4-5 days. All the case-patients reported having presented with reddening of eyes (Figure 1).

**Figure 1:**
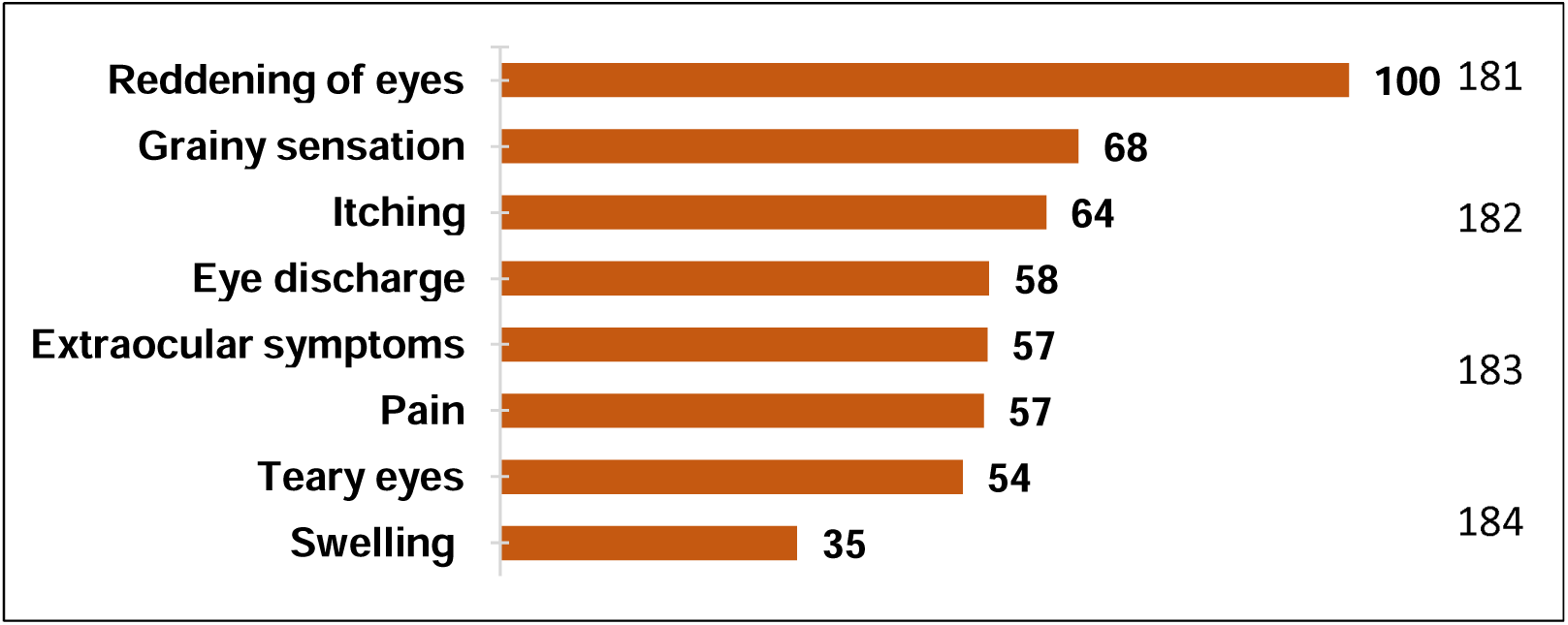
Distribution of symptoms among case-patients during a conjunctivitis outbreak, Luzira Prison, February – March 2024.

The overall AR was 23% (1,935/8,518), with males being more affected (96%, n=1,852) compared to females (4%, n=83). Murchison Bay Prison with 1,229 cases (AR: 41/100) was the most affected prison (Table 1).

**Table 1:** Attack rates by prison during an outbreak of conjunctivitis, Luzira Prison, Kampala, Uganda, February 1- April 2, 2024.

| Prison name | Cases | Total population | Attack rate (AR/100) |
| --- | --- | --- | --- |
| Murchison Bay Prison | 1,229 | 3,000 | 41 |
| Kampala Remand Prison | 610 | 1,835 | 33 |
| Murchison Bay Women's Prison | 83 | 670 | 12 |
| Luzira Upper Prison | 13 | 3,013 | 0.4 |
| **Total** | **1,935** | **8,518** | **23** |

This outbreak started with admission of infected inmates at Kampala Remand Prison from February 2024. Sharing of topical eye treatments between infected and non-infected inmates started on March 12, 2024. On March 15, there was a peak in the number of resident cases. This sharing of eye medication was discontinued on March 19, 2024 and inmates’ medication was administered separately by the “doctor ward”. Immediate isolation protocols for newly-admitted inmates were initiated thereafter. Infected inmates continued to be admitted into KRP beyond the time of the investigation (Figure 2), and the overall AR in Kampala Remand Prison was 33%.

**Figure 2:**
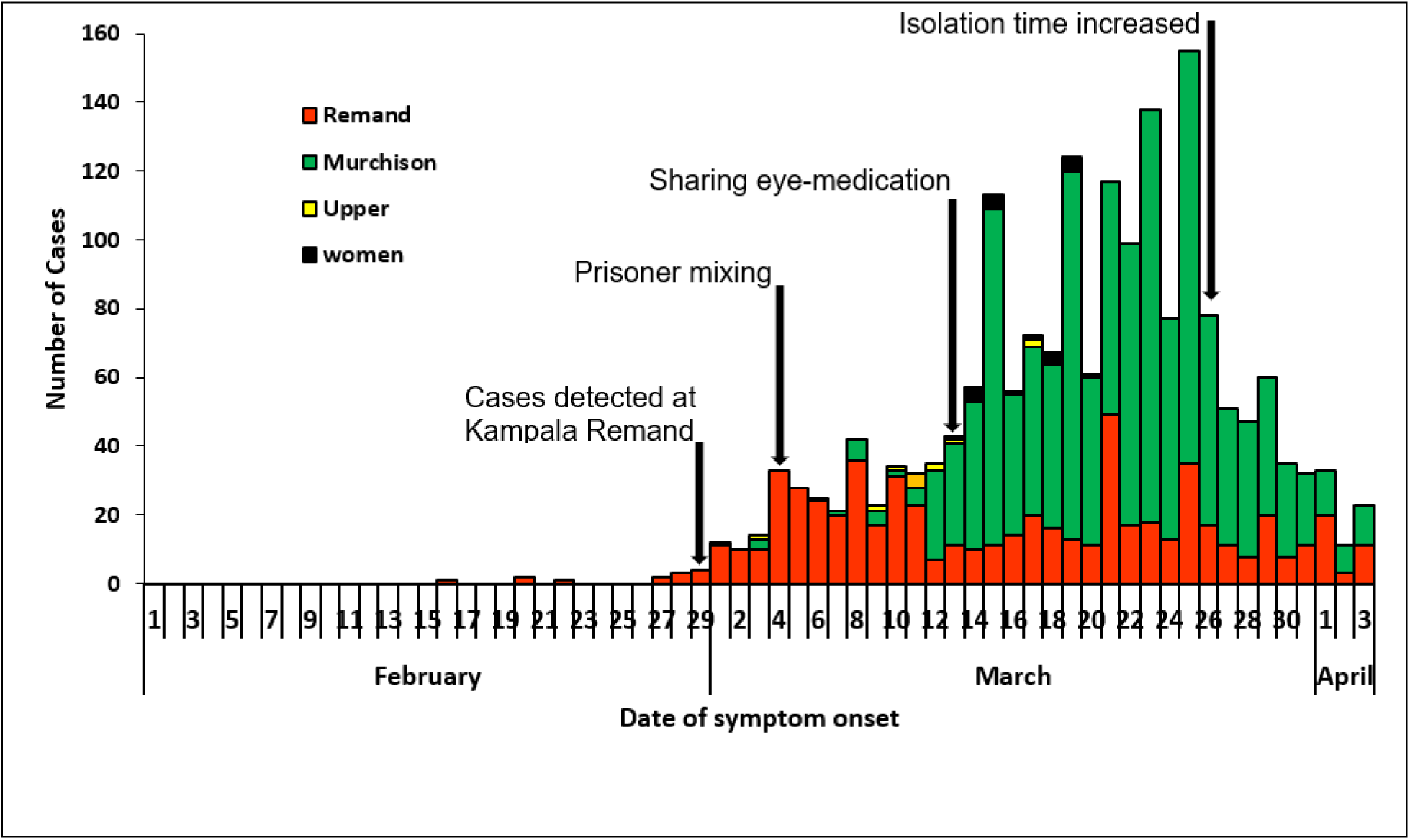
Distribution of conjunctivitis cases over time among case-patients in Luzira Prisons, February 1 – April 3, 2024 (n=1,935)

Some cases reported in Murchison Bay Prison involved prisoners referred to the Prison Hospital. These case-patients were promptly isolated on admission. On March 5, 2024, there was prisoner mixing during a plea-bargain meeting held within the confines of Kampala Remand Prison where some prisoners from Murchison Bay and Upper Prison attended (Figure 2).

Subsequently, more cases of conjunctivitis emerged among inmates at Murchison Bay Prison from March 6, 2024. As a preventive measure, there was distribution of tetracycline eye ointment (TEO) two days later. TEO served both as prophylaxis for those without symptoms and as a treatment for affected individuals. Despite these interventions, the number of cases steadily increased over time, marked by multiple peaks in incidence (Figure 2).

Starting from March 26, additional public health measures were implemented. These included prompt isolation of identified cases for at least five days, providing Continuous Medical Education (CME) sessions for healthcare workers and laboratory staff, conducting health talks for inmates. These public health measures preceded a noticeable decline in the number of reported cases (Figure 2).

### Women’s Prison Luzira

Cases were identified in March 2024 from inmates who attended court sessions and reported to have shared a bus with an infected inmate from Murchison Bay on her way to and from the court (Figure 2).

### Luzira Upper Prison

The data obtained from Luzira Upper Prison indicated that cases of conjunctivitis were routinely recorded in the health facility records.

Overall, the outbreak in Murchison Prison was preceded by that in Remand Prison, and by April 3, a total of 1,935 suspected cases had been reported to the prison health facilities from the 4 prisons.

### Laboratory findings

From the 10 samples collected and submitted for PCR testing, 4 (40%) were positive for Enterovirus Type C.

### Environmental and case management assessment findings

Kampala Remand Prison every day transported 150 – 200 inmates to attend different court sessions. In contrast, the maximum-security prison (Upper prison) transports an average of 80 inmates to courts daily and these courts are usually different from those which inmates from both Kampala remand and Murchison Bay prison go to.

Some inmates were released from court, while at the same time, new inmates were remanded. Murchison Bay Prison receives prisoners referred from other prisons for medical attention at Murchison Bay Prison Hospital. There was prisoner mixing during a plea bargain session at Remand prison where some inmates from MBP and LUP attended.

We observed the presence of tap and tank water at the prison main gate and at entrance to some prison wards. Where handwashing points existed outside the wards there was both soap and water, while those inside the wards had water pre-mixed with liquid soap.

In the earlier days of the outbreak, cases were isolated and treated. Later, the number of cases were so high that isolation was no longer possible. Identified cases were treated from the prison wards where inmates resided. Treatment was being administered topically by the “doctor ward” using their hands to all inmates in spite of the disease status of the inmates. This was not observed in both the Upper prison and the women’s prison. By this time, only those that had severe eye discharges were isolated. Isolation initially lasted only up to 3 days in Murchison Bay prison. However, in LUP, isolation was sustained until prisoners recovered fully from the infection.

### Hypothesis generation findings

Based on the descriptive epidemiology, laboratory, and the environmental assessment findings, we hypothesized that sleeping next to some sick and sharing eye medication was associated with propagation of the outbreak among inmates. Poor hand hygiene was associated with increased transmission of conjunctivitis among inmates (Table 2).

**Table 2:** Exposures among 200 cases during a conjunctivitis outbreak, Luzira prisons, Uganda, March–April 3, 2024.

| Factor | Cases (n=200) | % |
| --- | --- | --- |
| Washed face daily | 195 | 98 |
| Slept next to someone sick | 175 | 88 |
| Washed hands with soap | 139 | 70 |
| Moved out of prison | 114 | 57 |
| Shared blankets | 63 | 32 |
| Shared eye medication | 51 | 26 |
| Received visitors | 34 | 17 |

### Case control investigation findings

We enrolled 200 controls for the case-control study. At bi-variate level, sharing eye medication (cOR:3.6, 95%CI=2.0-6.6), sleeping next to someone sick (cOR:2.2, 95%CI=1.3-3.7), and reduced hand-washing frequency (cOR:5.2, 95%CI=3.2-8.3),) were associated with conjunctivitis. Results from multivariable analysis indicated that sharing of eye medication (aOR:4.7, 95%CI=2.5-8.7), and reduced frequency of handwashing (aOR:5.8, 95%CI=3.5-9.6) were significantly associated with acquiring the infection among inmates (Table 3).

**Table 3:** Factors associated with conjunctivitis among case-persons during an outbreak, Luzira prisons, Uganda, March–April 3, 2024.

|  | Cases |  | Controls |  |  |  |  |  |
| --- | --- | --- | --- | --- | --- | --- | --- | --- |
| Variable | n=200 | % | n=200 | % | cOR | 95% CI | aOR | 95% CI |
| **Sharing eye medication** |  |  |  |  |  |  |  |  |
| No | 149 | 75 | 183 | 91 | Ref | Ref | Ref | Ref |
| Yes | 51 | 25 | 17 | 9 | 3.6 | 2.0-6.6 | 5.2 | 2.8-9.5 |
| **Sleeping next to someone sick** |  |  |  |  |  |  |  |  |
| No | 26 | 13 | 49 | 25 | Ref | Ref | Ref | Ref |
| Yes | 175 | 87 | 150 | 75 | 2.2 | 1.3-3.7 | 1.8 | 0.9-3.1 |
| **Received visitors** |  |  |  |  |  |  |  |  |
| No | 167 | 83 | 131 | 66 | Ref | Ref |  |  |
| Yes | 34 | 17 | 68 | 34 | 0.4 | 0.2-0.6 |  |  |
| **Moved out of prison** |  |  |  |  |  |  |  |  |
| No | 87 | 43 | 69 | 35 | Ref | Ref |  |  |
| Yes | 114 | 57 | 130 | 65 | 0.7 | 0.5-1.1 |  |  |
| **Shared blankets** |  |  |  |  |  |  |  |  |
| No | 138 | 69 | 112 | 56 | Ref | Ref |  |  |
| Yes | 63 | 31 | 87 | 44 | 0.6 | 0.4-0.9 |  |  |
| **Did not Wash hands with soap and water** |  |  |  |  |  |  |  |  |
| Yes | 62 | 31 | 8 | 4 | Ref | Ref | Ref | Ref |
| No | 139 | 69 | 191 | 96 | 10.6 | 4.9-22.9 | 13.9 | 6.4-30.5 |
| **Frequency of washing hands** |  |  |  |  |  |  |  |  |
| Frequently/Always | 105 | 48 | 169 | 85 | Ref | Ref | Ref | Ref |
| Rarely/ Occasionally | 96 | 52 | 30 | 15 | 5.2 | 3.2-8.3 | 5.8 | 3.5-9.6 |

**cOR: Crude odds ratio; aOR: Adjusted odds ratio; Ref: Reference category**

## Discussion

We investigated an outbreak of conjunctivitis within a Uganda prison setting, to assess the magnitude and risk factors associated with the outbreak, and recommend control measures. The outbreak primarily affected Kampala Remand Prison (KRP) and Murchison Bay Prison (MBP), where the attack rates were notably higher compared to Luzira Upper Prison (LUP) and Luzira Women Prison (LWP). Most cases were mild, with rapid resolution within 4-5 days, and no fatalities were reported. Laboratory testing confirmed Enterovirus Type C as the cause. Risk factors, including sharing eye medication and infrequent handwashing, were strongly associated with increased infection. Interactions between inmates from different prisons and suboptimal isolation of symptomatic individuals likely contributed to the spread. These findings highlight the need for improved hygiene practices and stronger isolation protocols in high-density, closed environments like prisons.

The conjunctivitis outbreak was attributed to Enterovirus Type C, which aligns with findings from other outbreak investigations indicating that enteroviruses are the primary cause of most viral conjunctivitis outbreaks (1–3). Symptoms resolved within 4-5 days, with no severe complications reported. These results are consistent with previous studies, which indicate that the duration of illness typically ranges from 2 to 7 days without major complications (7, 15–17).

Murchison Bay Prison was the most affected prison. The variation in attack rates can be attributed to the unique operational characteristics of each prison. For example, KRP experiences a high turnover of inmates, with new arrivals from police cells and courts, which likely contributed to the initial case and subsequent spread of conjunctivitis. The outbreak at MBP followed that at KRP, likely due to prisoner mixing. Inmates from MBP who attended a plea-bargain session at KRP were among the first cases. These meetings, which bring together inmates from different prisons, provide a conducive environment for disease transmission. To prevent future outbreaks, prison authorities should consider implementing screening measures to exclude affected individuals from such gatherings. Additionally, organizing separate plea-bargain sessions for different prisons could help reduce the risk of cross-prison transmission [10].

The administration of topical eye medications may have inadvertently contributed to the propagation of the outbreak among inmates. Prison ward leaders, referred to as ‘doctor wards,’ were administering eye medications to both infected and non-infected inmates using the same tubes and initially applying TEO directly with their bare hands. This direct contact likely facilitated the transmission of the infection from infected to uninfected inmates. Previous studies have shown that sharing eye medications can promote the spread of conjunctivitis, with additional evidence highlighting an increased risk associated with such practices (16). To mitigate the rapid spread of infection within prison settings, it is essential to reduce contact between infected and non-infected inmates. Implementing screening protocols, ensuring prompt isolation, and providing individualized treatment for cases could significantly help in controlling outbreaks.

Initially, all identified case-persons were isolated; however, as the number of cases increased significantly, isolating every affected individual became impractical due to space constraints. The prison was already operating at approximately 300% of its original capacity. As a result, isolation was limited to symptomatic individuals, with a maximum duration of 3 days. Previous studies have demonstrated that isolation of infected individuals is an effective control measure (18). Based on our findings, we recommend extending the isolation period to the recommended 5 days during future outbreaks to reduce transmission risk among inmates.

After the isolation period, symptomatic inmates were returned to their respective wards, and the remaining cases were managed within overcrowded conditions, leading to inevitable close contact between infected and non-infected individuals. Close proximity to infected inmates and the subsequent sharing of clothes and blankets were associated with a higher likelihood of developing conjunctivitis. This finding aligns with studies from Brazil, the USA, and China, where conjunctivitis spread through the sharing of fomites among infected individuals (5, 19).

Frequent hand washing with soap and water prevented infection spread, consistent with findings from previous studies elsewhere (15). During conjunctivitis outbreaks, prison authorities should prioritize the availability of handwashing facilities to promote better hygiene practices and reduce the risk of transmission.

### Study limitations

A key limitation of this study is its scope, as we focused exclusively on the outbreak within Luzira Prisons, which limited our ability to assess the broader evolution and extent of the outbreak across other prisons in Uganda. As a result, potential clusters in other facilities or communities, such as schools, were not explored. This restriction prevented us from determining whether these clusters were epidemiologically linked to the Luzira prison outbreak. The lack of these broader epidemiological insights limited our ability to develop more comprehensive, evidence-based control measures.

## Conclusion

Our investigation identified Enterovirus type C as the causative agent of the conjunctivitis outbreak, which had varying impacts across different prison facilities, with no fatalities reported. The initial transmission was linked to the admission of an infected inmate, with further spread attributed to inmate interactions during a mass meeting (plea bargain sessions). Contributing risk factors to the transmission included sharing of topical eye medications, overcrowding, close contact between symptomatic and asymptomatic inmates, and inadequate isolation practices. However, frequent handwashing was associated with a reduced risk of infection. We recommend strengthening infection prevention and control measures, including strict isolation of affected individuals, individualized eye medication, and enhanced hand hygiene practices to mitigate the risk of future outbreaks.

### Public health actions

During the investigation, we conducted sensitization sessions for inmates, emphasizing the importance of proper hand hygiene. We also trained the “doctor ward” staff on the correct administration of eye ointments and drops, highlighting the need for prompt reporting and referral of suspected cases to health facilities. Additionally, continuing medical education (CME) sessions were held for healthcare workers, and clinical and laboratory staff received training on sample collection, storage, and referral procedures. These were all in based on evidence-based management strategies from other studies In collaboration with the prison administration and health facility staff, we implemented immediate control measures. All identified cases during case-finding were referred to the prison health facility for further management. Following the dissemination of our findings, the isolation period was extended from 3 days to 5-7 days (20).

We conducted an assessment of the prison laboratory’s outbreak readiness and provided recommendations for improvement to the prison authorities and relevant partners. A preliminary report of the findings was presented to the Public Health Commission of Uganda Prison Services for further review and action.

## Data Availability

The datasets upon which our findings are based belong to the Uganda Public Health Fellowship Program. For confidentiality reasons, the datasets are not publicly available. The datasets can be availed upon reasonable request from the corresponding author with permission from the Uganda Public Health Fellowship Program.

https://www.msdmanuals.com/professional/eye-disorders/conjunctival-and-scleral-disorders/overview-of-conjunctivitis

## Author Contribution

Hannington Katumba led the investigation and drafted the manuscript. Hannington Katumba, Charity Mutesi, Emmanuel Mfitundinda, Joanita Nalwanga, Owens Joyce Kobusngye, Daniel Wenani, Loryndah Olive Namakula, Emmanuel Okello Okiror, Janet Lubega Kobusinge, Gertrude Abbo, Annet Mary Namusisi, Bridget Ainembabazi, Patrick Kwizera, Wilfred Opeli, Winnie Agwang, Esther Nabatta, Tract Maureen Rutogire, Ritah Namusoosa and Samuel Lugwana, investigated the outbreak and participated in the design of data collection tools, data collection, entry and cleaning, analysis, and review of the manuscript draft. Richard Migisha, Benon Kwesiga and Sam Gidudu supervised the outbreak investigation and reviewed the manuscript for intellectual content. Richard Migisha, Benon Kwesiga, Lillian Bulage, and Alex Riolexus Ario reviewed manuscript draft for intellectual content and scientific integrity.

## Acknowledgements

We extend our sincere gratitude to the Kampala Capital City Authority/Kampala Metropolitan Area Emergency Operations Center team for their prompt notification to the Ministry of Health, facilitating swift response measures. We also wish to express our appreciation to Dr Aida Ajambo and the entire Public Health Commission of the Uganda Prison Service for their exemplary coordination efforts and responsive administrative support, enabling us to access the inmates effectively.

## Funding and disclaimer

This study was supported by the President’s Emergency Plan for AIDS Relief (PEPFAR) through the United States Centers for Disease Control and Prevention Cooperative Agreement number GH001353-01 through Makerere University School of Public Health to the Uganda Public Health Fellowship Program, Ministry of Health. The contents of this manuscript are solely the responsibility of the authors and do not necessarily represent the official views of the US Centers for Disease Control and Prevention and the Department of Health and Human Services, Makerere University School of Public Health, or the Uganda Ministry of Health.

